# The Covid-19 infection in Italy: a statistical study of an abnormally severe disease

**DOI:** 10.1101/2020.03.28.20046243

**Authors:** Giuseppe De Natale, Valerio Ricciardi, Gabriele De Luca, Dario De Natale, Giovanni Di Meglio, Antonio Ferragamo, Vito Marchitelli, Andrea Piccolo, Antonio Scala, Renato Somma, Emanuele Spina, Claudia Troise

## Abstract

We statistically investigate the Coronavirus Disease 19 (hereinafter Covid-19) epidemics, which is particularly invasive in Italy. We show that the high apparent mortality (or Case Fatality Ratio, CFR) observed in Italy, as compared with other countries, is likely biased by a strong underestimation of infected cases. To give a more realistic estimate of the mortality of Covid-19, we use the most recent estimates of the IFR (Infection Fatality Ratio) of epidemic, based on the minimum observed CFR, and furthermore analyse data obtained from the ship Diamond Princess, a good representation of a ‘laboratory’ case-study from an isolated system in which all the people have been tested. From such analyses we derive more realistic estimates of the real extension of the infection, as well as more accurate indicators of how fast the infection propagates. We then point out from the various explanations proposed, the dominant factors causing such an abnormal seriousness of the disease in Italy. Finally, we use the deceased data, the only ones estimated to be reliable enough, to predict the total number of infected people and the interval of time when the infection in Italy could stop.

## Introduction

The infection of Covid-19, recently declared pandemic by WHO, represents perhaps one of the most serious worldwide emergencies, potentially able to destroy social order and economies and to deeply change our lifestyle in the near future. Since the first appearance of this new Coronavirus (SARS-CoV-2) (Worldometers, 2020), however, the Covid-19 infection has been treated with very mixed feelings: from just a disease a little more serious than a seasonal flu, to a very severe and troubling infection. The epidemics was firstly detected in China, in the city of Wuhan, at the end of December 2019. After some interlocutory days, the government of China showed serious concern and implemented very severe measures in the Hubei province, the centre of the epidemics, to contain the epidemic spreading of the infection. About 45 days after the first detection (mid- February 2020), the epidemics started to seriously affect several other countries (South Korea among the firsts, being a rather natural candidate that borders with China). Since the end of February, it flared up in Italy and Iran for less clear reasons and, since mid-March 2020, the epidemics has spread all over Europe, in the USA, and many other countries (Worldometers, 2020). Here we want to focus on the Covid-19 epidemic in Italy that shows some peculiar features, distinguishing its evolution from the one observed in other countries. The epidemic appears very aggressive, both in terms of spread rate and mortality, which are however very uncertain parameters for this new virus. In Italy, the infection is mainly focused in the Lombardia Region and the area around the Po river. The most affected regions are Lombardia, Emilia-Romagna and Veneto, which also represent the richest and more productive parts of Italy. In Italy the infection grew very fast, overcoming South Korea in the number of infected people as early as in the beginning of March 2020, today (March 30^th^, 2020) reaching 101739 total infections. Moreover, it showed an average Case Fatality Ratio (CFR) over 11%, well above any other country and more than double with respect to the Hubei Region in China, where the new virus first appeared, and where CFR was significantly higher than in other parts of the China and higher than several countries in the world.

In this paper, we will show statistical analyses of data associated to the Italian Covid-19 epidemics. The aim is to estimate a possible slowdown of the infection and also to verify our statistical predictions in relation with the severe containment measures taken by the Italian Government. We discuss alternative explanations for the very high CFR observed. To confront with the important problem of determining the true mortality (Infection Fatality Ratio, IFR) of the epidemics, we use the study of an isolated, well-calibrated test case, represented by the infection spread on the Diamond Princess cruise ship. Mortality (IFR) estimates obtained within the Diamond Princess are an ‘unbiased’ value, not affected by the underestimation of the number of infected people; also IFR values computed by the University of Oxford (Oke and Henegan, 2020) were checked. Using various IFR estimations, we predict a much larger number of infected people in Italy with respect to the official one, suggesting that the CFR highly overestimates true mortality. We also discuss the likelihood of alternative hypotheses, often claimed to explain the high impact of Covid-19 in Italy, based on the influence of old average age of population, high antibiotic resistance, high number of smokers, pollution in the Po plain (e.g. Oke and Henegan, 2020). Finally, we identify the best data set to analyse the statistical evolution of the epidemics in Italy, also comparing it with its prototypical behaviour obtained from the China dataset, to try to forecast the time of saturation of the infection.

### The Covid-19 in Italy

The Covid-19 epidemic in Italy presents some peculiarities that make it very intriguing to analyse and understand. Initially it was thought that two independent focuses started in Codogno (15962 inhabitants, Lodi province, Lombardia Region) and Vo’ Euganeo (3416 inhabitants, Padua province, Veneto Region) towns, but it is now generally understood that the virus started circulating earlier in the whole north of Italy. The epidemic rapidly blew up all over Italy, but particularly around the Po Valley, in Lombardia, Veneto and Emilia-Romagna. These Regions are the richest ones in Italy, for their industries, agriculture in the Po Valley and international commerce. The Lombardia and the Po Valley, the most hit by the infection, are also the most polluted areas in Italy and probably also the most polluted in the whole Europe by the fine air particulate matter (PM10, PM2.5) and Ozone (Martuzzi et al., 2006; Stafoggia et al., 2009).

The beginning and evolution of Covid-19 infection in Italy, and more specifically in Lombardia Region, is from many points of view anomalous and highly lethal, with respect to all other countries worldwide, including China where the infection was born. Although all the media (and also many specialists interviewed by media) highlight the velocity of infection in Italy as ‘exponential’, the number of infected people has never followed an exponential distribution, except in the very first few days. Figure 1a shows the number of recorded infections as a function of time (in days since February 24^th^ 2020) in a semi-logarithmic scale. It is evident that the distribution is markedly different from a straight line, typical of an exponential distribution in such a scale, and it’s rather well fitted by a cubic polynomial, which is much slower than an exponential. The cubic polynomial fit and the exponential one present in the figure are both carried out using the data up to 18 days from the date of acknowledgment of the first cases (from 24^th^ February to 12^th^ March); starting from that date the cubic fit scores, according to an Akaike information criterion (AIC) test, better than the exponential one. In the figure we also show the fit obtained from a logistic function for comparison; logistic fit scores (AIC test; Akaike, 1974) become better than the exponential one from 12^th^ March and definevely better of the cubic one after 25^th^ March. Figure 1b shows the same quantity in a linear scale, together with the three mentioned fitting functions.

**Figure 1.**
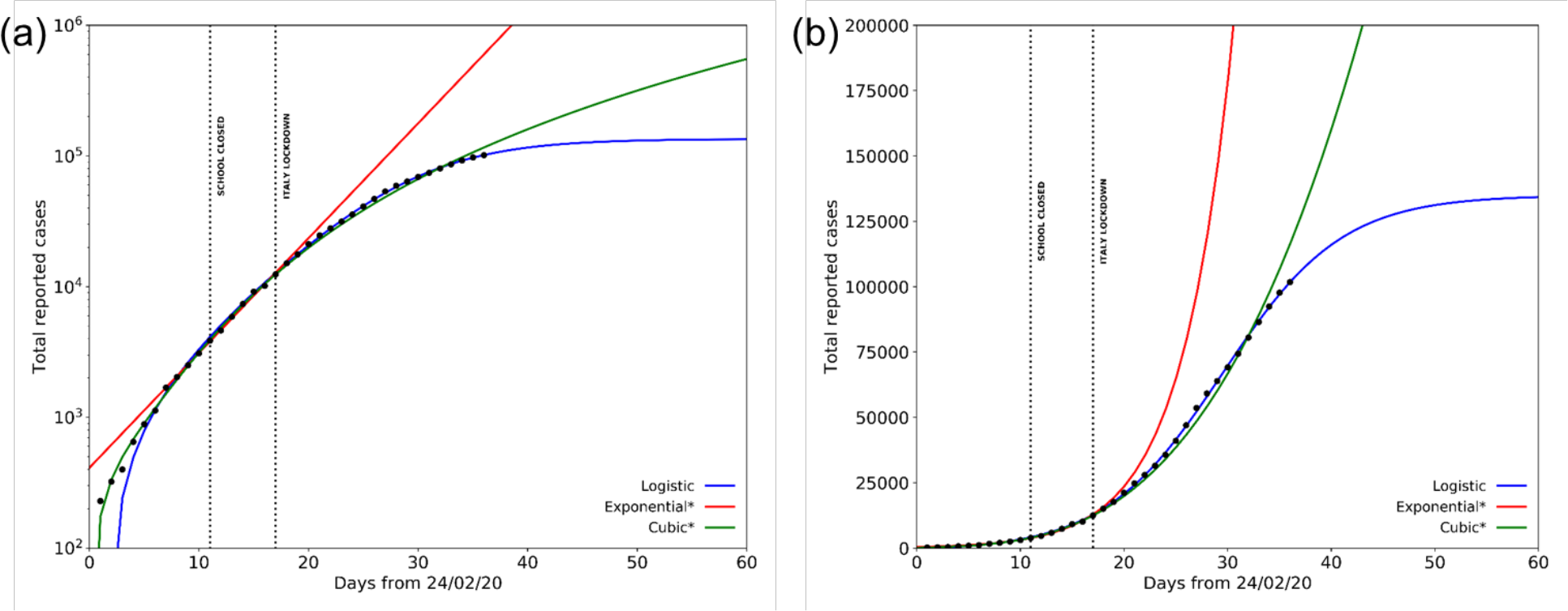
Total Covid-19 reported cases in Italy from February, 24^th^ to March, 30^th^ according to Protezione Civile (black dots) with the logistic (blue solid line), exponential (red solid line) and cubic (green solid line). Dotted black vertical lines mark the dates of Italian school lockdown and Italy total lockdown; the asterisk indicates that the exponential and cubic fit are based on data til March, 12^th^: score from AIC test (not reported) on the logistic, cubic and exponential fit shows higher reliability of the first two after this date, and for the logistic against the cubic after 25^th^ March a) fits obtained from the data in semi-logarithmic scale; B) same data and fits shown in linear scale. Fit parameters: Logistic (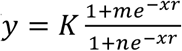; *K* = 135 (± 2) · 10^3^,*m* = −1.4 ±0.2, *n* = 170±9,*r*=0.174±0.003); Exponential (*y* = *Ae*^*Bx*;^*A* =410 ±30,*B*=0.2 ±0.01;*Cubic*= *a*+*bx*+*cx*^2^+*dx*^3^;*a* = −40± 20,*b*=36±8,*c*= −6.7±0.9,*d*=0.44±0.03.

Although the number of infected people is the main parameter taken into account by the authorities, and the most highlighted by media, its real value is largely uncertain and surely underestimated. In fact, it critically depends from the number of laboratory tests made on people to ascertain the infection, which is anyway limited and very small as compared with the number of inhabitants. Furthermore, the procedure to test people are highly variable within the different Regions of Italy and have changed over time in the last weeks; because of this inconsistency, this number is statistically very inhomogeneous and unfit to to interpret the actual evolution of the infection.

The number of tests in Italy, highly fluctuating but generally increasing in time except in the last days, ranged from about 2427 (February 27^th^) to 26336 (March 21^st^), and decreased again to 25180 (March 22^nd^) and 17066 (March 23^rd^). More recently, it reached a peak value of 36615 on March 26^th^ but then it decreased in the next days (Il Sole 24 Ore, 2020). A very important quantity, namely the CFR, defined as the ratio of number of people deceased divided by number of total recorded infections, is extremely high in Italy (about 11%). Such a high value is dominated by the mortality in Lombardia, where about 50% of all the Italian infections have been recorded, with a CFR of about 16%. CFR is a generally overestimated value of the true mortality (IFR), given the likely underestimation of the real number of infection cases (i.e. including asymptomatic and pauci- symptomatic cases, which are easily overlooked by the small number of tests). IFR is the parameter which measure the percentage of deceases over the total population infected (including the generally unknown number of non recorded cases). Table 1 (Oke and Henegan, 2020) reports the number of infections, deaths and CFR observed in several countries in the world as of March 30^th^, 2020. It is possible to note that the mortality rate for different countries is very variable, going from a minimum of 0.4% (i.e. Australia, Israel) to a maximum of 11.3% in Italy. Several observations are however around 1%-2% (Germany, USA, Austria, Portugal, Ireland, etc.). It appears evident, therefore, that the CFR of Italy, and even more of Lombardia Region, is absolutely extreme when compared to any other country. This CFR is more than double of the CFR obtained in China, where the epidemic first appeared. Therefore, besides CFR, it is important to obtain a reasonable estimation of the IFR already during the epidemic spread, to understand the real hazard of the disease and/or to determine the real amount of infected people.

**Table 1.**
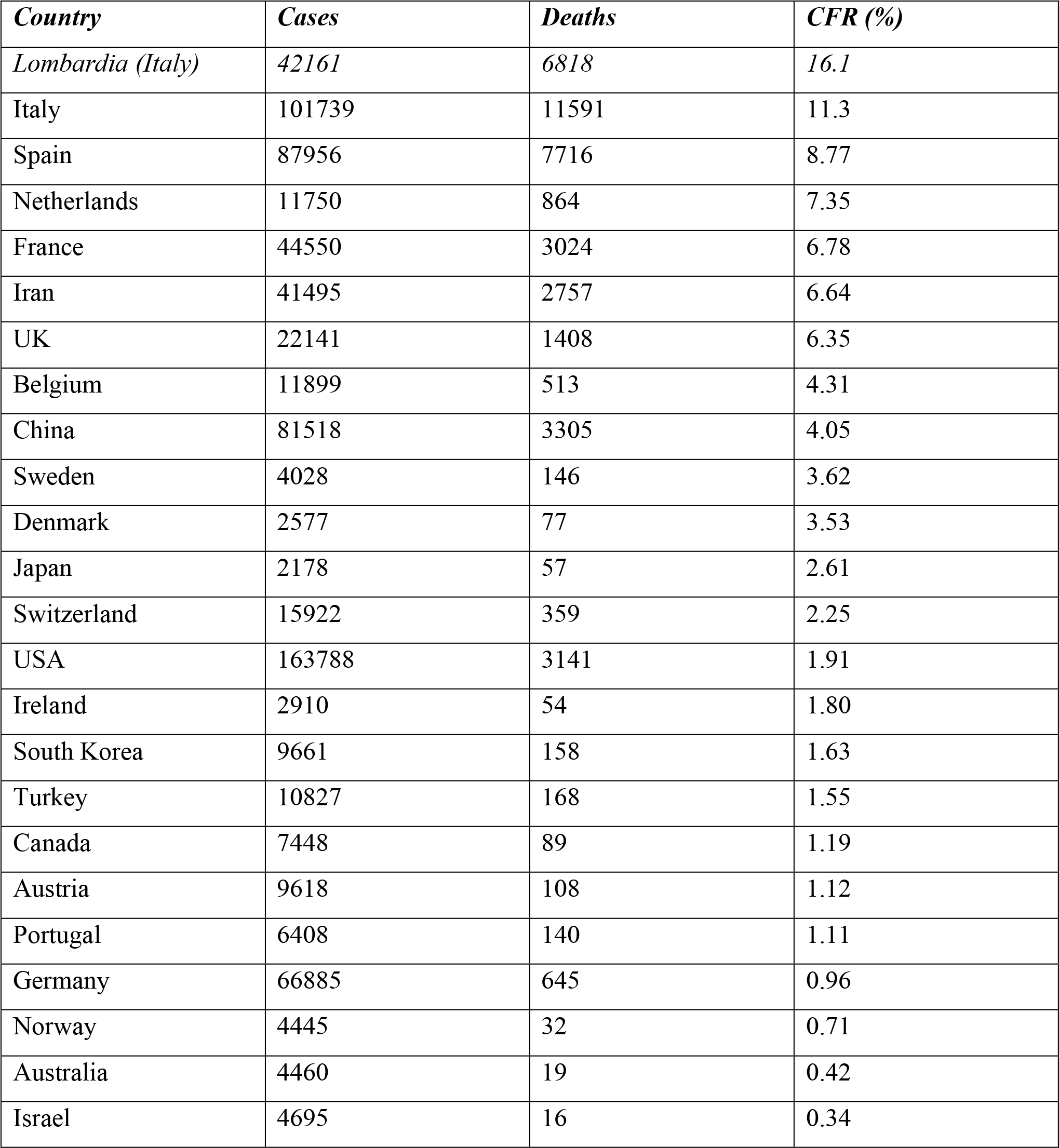
Numbers of recorded infected people and deaths in several countries, as on March 30th 2020. Also indicated is the CFR (Case Fatality Ratio) defined as the ratio between the number of deaths and the number of recorded cases.

### The IFR for Covid-19 and the mortality in Italy

The problem of unbiased estimation of the IFR for Covid-19 is complicated by the complex laboratory procedures that are necessary to identify the infected cases, ultimately limiting the total number of tested individuals. Since the IFR is the ratio between the number of deceased due to the disease and the total number of infected, the CFR, which is computed on the number of ‘known’ infected is an upper limit of IFR. Normally, CFR (and hence IFR) should be determined at the end of epidemic, because decease occurs at the end of the epidemic cycle, delayed with respect to the infection. Usually, at the very beginning of an epidemic, the CFR can be underestimated just because the outcome of the disease (recovered or deceased) has not yet been reached. For Covid-19, however, it was noted in China (and in South Korea too) that CFR was generally higher in the first phase of the infection spreading. A further problem is that the amount of underestimated infected cases is generally variable and could become progressively more critical as the real number of infected rapidly rises with time, while the number of tests remains stationary. Furhtermore, mass- testing in the village of Vo’ Euganeo suggested that the number of asymptomatic persons infected by the Covid-19 is at least 50% (La Repubblica, 2020). From Table 1 we observe that each country has a very variable CFR associated to the Covid-19 infection. Since the IFR is intrinsically overestimated by the CFR, presuming the virus strain is the same in every country, we can assume the minimum value of the observed CFR as the minimum upper limit for IFR. Following this procedure, the University of Oxford used one of the minimum CFR values (around 0.4%), namely from Israel, Australia and recently also Iceland, and to properly account for the undetected asymptomatics, halved it to obtain an IFR = 0.2% (Oke and Henegan, 2020). On the other hand, Italian CFR is very similar to what is observed in hospital casistic (11%) (Chen N. et al, Lancet 2020), which attest the attitude to test especially in-hospital and severe cases, being unaware of mild and asyntomatic patients.

For the Covid-19 epidemics, moreover, there is an independent way to obtain a rather unbiased estimate of IFR using the only ‘laboratory-like’ case study: the Diamond Princess, the cruise ship anchored in the port of Yokohama from February 4^th^ to March 2^nd^. Here, all the 3711 passengers and crew were tested and 712 (19.2%) persons had positive test results. Of these, 331 (46.5%) were asymptomatic at the time of testing, a similar value to the one obtained in Vo’ Euganeo. Among 381 symptomatic patients, 37 (9.7%) required intensive care, and 9 died (Moriarty L. F. et al., 2020). Because the Diamond Princess is a perfectly isolated case in which all the people have been tested, the total number of infected is perfectly known and we can assume that IFR=CFR. The crude IFR computed from the Diamond Princess is then 1.3%. Because this number is statistically affected by a rather large uncertainty due to the small number of deceased, we can then assume that the IFR of Covid-19 is less than 1.3%; indeed, the observation of a CFR significantly smaller than 1% in some countries (i.e. Australia, Norway, Israel) indicate that lower values are more probable than larger ones. In the light of such derived mortality range, it appears very abnormal the observed CFR in Italy (11.3%), and even more the CFR in the Lombardia Region (16.1%).

In the following, we will try to point out the possible explanations for such an extremely anomalous outcome. We will, again, assume that the virus strain is the same in every country, since there are not contrary evidences till now. The first, more obvious reason to explain such a high mortality, is to hypothesize that the number of infected cases is substantially underestimated. A clear sign of this is the fact that mortality increases during time, from around 2% at the beginning (February 20th) to 11% on March 30^th^. During this period the number of detected cases rose from few units to around 100000, whereas the daily number of laboratory tests changed from few hundreds to about 30000.

Besides the CFR, which is only linked to the recorded infection cases, what is really interesting to understand is a ‘local’ IFR estimation for Italy, in order to derive the real total number of infected people. Lacking evidence for the existence of a different virus strain, more aggressive and lethal in Italy, we firstly assume that IFR is the same as in other places, and a distinctive trait of this epidemic. We could then choose between the University of Oxford estimate of IFR=0.2% or the Diamond Princess laboratory-like estimation of IFR=1.3%. If the extreme mortality observed would be only due to the underestimation of the number of infected people, assuming a ‘true’ IFR ranging between 0.2% and 1.3%, to correct for the Italian 11.3% CFR, we should multiply, respectively, by 56.5 or by 8.7 the official number of infected cases (101739 on March 30^th^); the resulting number would then range between about 885000 and 5.7 million infected people.

We cannot exclude, however, the true IFR in Italy to be significantly higher than in other countries because of several concurrent reasons. For instance, the higher average age of Italian people has often been indicated as a possible explanation for the high Covid-19 CFR. Among all the countries of the world, Italy is actually in the second position for higher average age; however, the first position (oldest population) is occupied by Japan, which showed a very low number of infections and CFR (see Table 1) so that this possibility appears unlikely.

Another possible cause of comorbidity could be related to the high level of pollution in Lombardia Region that is likely the most polluted region in Europe by fine particulate (PM10, PM2.5) and Ozone. As shown in several papers (e.g. Chen et al., 2010; Ye et al., 2016; Chen et al., 2017; Setti et al., 2020) there is a correlation between the diffusion of viruses and the pollution by fine particulate. Furthermore, exposition to fine powders contributes to enhance the severity for respiratory viral infections (Dominici et al., 2006; Ciencewicki and Jaspers, 2007). The incidence of fine particulate pollution could hence, in principle, be one of the reasons for the high mortality rate observed in Lombardia (and partially in Emilia-Romagna, around the Po Valley). Although an effect of this kind of pollution in amplifying the mortality observed for a severe pulmonary disease seems reasonable, it is actually very difficult to quantify its incidence. It is also very difficult to believe it can be so strong with respect to other highly industrialized areas of Europe (e.g. Germany, France, Netherlands, etc.). Moreover, a recent document issued by the Italian Aerosol Society, signed by about 60 scientists of various disciplines, rejects various hypotheses (Setti et al., 2020), pointing out that there is no clear evidence for a correlation between fine particulate and Covid-19 disease amplification (Contini et al., 2020).

Other tentative explanations for the extreme mortality could be the high number of smokers in Italy and the antibiotic-resistance of Italian people. Regarding the percentage of smoking people, however, Italy’s 23% is lower than the European average, 29% (WHO, 2016); regarding the antibiotic-resistance, on the contrary, Italy has actually the most critical position in Europe. Among about 33000 yearly deceases in EU due to antibiotic-resistant bacteria, about 10000 occur in Italy alone (ISS, 2019). Since generally therapies used against the SARS-CoV-2 involve one or more antibiotics, this issue could, in principle, result in higher mortality. Also in this case, however, it is not easy to quantify this effect, which however appears marginal in seasonal flu, since in this case CFR for Italy does not differ too much with respect to other European countries (ISS, 2019). Moreover, Covid-19 antibiotics administration aims to avoid bacterial superinfection, whose mortality is negligible; so even if there was a more pronounced antibiotic resistance and this mechanism would be implied in higher mortality, this would probably account for only a very little part of the deaths. The remaining possibility is that the healthcare was unprepared for such an emergency due to a respiratory syndrome; actually, we just note that in Italy (60 million people), there were, before Covid-19 epidemic, about 5090 places in Intensive Care Units (ICU); for comparison in Germany (82 million people) such places were 28000. Actually, in terms of number of ICU divided by population, Italy occupies the 19th position among 23 European Countries. We observe indeed high pressure on ICU by severe/critical cases, mainly in Lombardia, where the number of ICU before the crisis was 900; this number increased to over 1000 during the last weeks, but, as of March 30^th^, the patients hosted in ICU are more than 1300. Indeed, already on March 14^th^, the Government of Lombardia declared that available ICU were almost over. Another indication that something went wrong during the first phase of management of the infection by the Lombardia Hospitals is the very high number (6414) of infected medical staff (ISS daily Info, March 30^th^). So, the Hospitals could have been the most effective carriers for the epidemic in the first phase in Lombardia, when a very fast increase was observed.

### Possible forecast of future behaviour of infection

In order to forecast the evolution and the end of the Covid-19 epidemic in Italy, we could in principle use three kinds of data. The most obvious would be the daily infection data. However, such data are particularly unreliable, because too much dependent from the daily number of tests. They are generally very variable and inhomogeneous, both from a Region to another, and also in time. Since, as we noted, the real number of infected people is likely much higher (orders of magnitude) with respect to the sampled one, the inhomogeneous sampling can strongly condition the number of infections, making them not useful for a statistical study. Another possible indicator of the epidemic evolution is the number of people in ICU. Contrarily to the number of infected, the number of people in ICU should be objective, because who has serious breathing problems must necessarily be hospitalized in ICU. However, in this phase of the epidemic crisis, this number has two major problems: the first one is that ICU places are mostly full, at least in Lombardia which dominates the Italian statistics, so not all people requiring them can be allocated. The second problem is that the daily numbers given by Italian Civil Protection just mention the total number of people hosted in ICUs in that day, and not the daily incremental number. Hence, it is not possible to know the real cumulative number of people hosted in ICUs, because we don’t know how many people each day came out from them, due to recovering or death.

The only quantity which has a rather rigorous statistical meaning, then, is the daily cumulative number of deceases. We then choose to use this number in order to statistically analyse the evolution of epidemic and to predict its end. Obviously, since we are particularly interested in determining the time at which the epidemic ends and the total number of infected people cumulated at the end of the epidemic, we have to correctly consider the relation of the daily cumulative number of deceases with the cumulative number of infected people. The number of deceased people is linked to the number of infected one by the IFR=D/I (D= deceased, I=infected); then, correcting the number of deceases for the constant factor represented by the inverse of IFR (I/D) gives the number of infected people. However, we must also consider that infection and decease are just the two temporal limits of the disease: it starts with infection, proceeds with the symptoms, and then ends with one of the two possibilities: recovering or death. To account for this, we must consider the shift in time between the infection and the decease. For Covid-19, it has been estimated that the average time from infection to death is 16 days, with this value being about the median in the interval of confidence [13.1 – 17.7] (Jung et al., 2020). With such relations in mind between infected and deceased people, we fit the decease data to a logistic function of the general form: 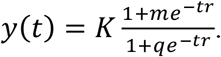

To validate the use of this function, in Figure 2 we show the fit made using a logistic function, to the epidemic curve of the cumulative daily number of cases (infections) reported in the whole China, Hubei province and China without Hubei province. It is clear that, after the epidemic spread is over, the whole epidemic behaviour from the start to the end can be well described by the logistic function.

**Figure 2.**
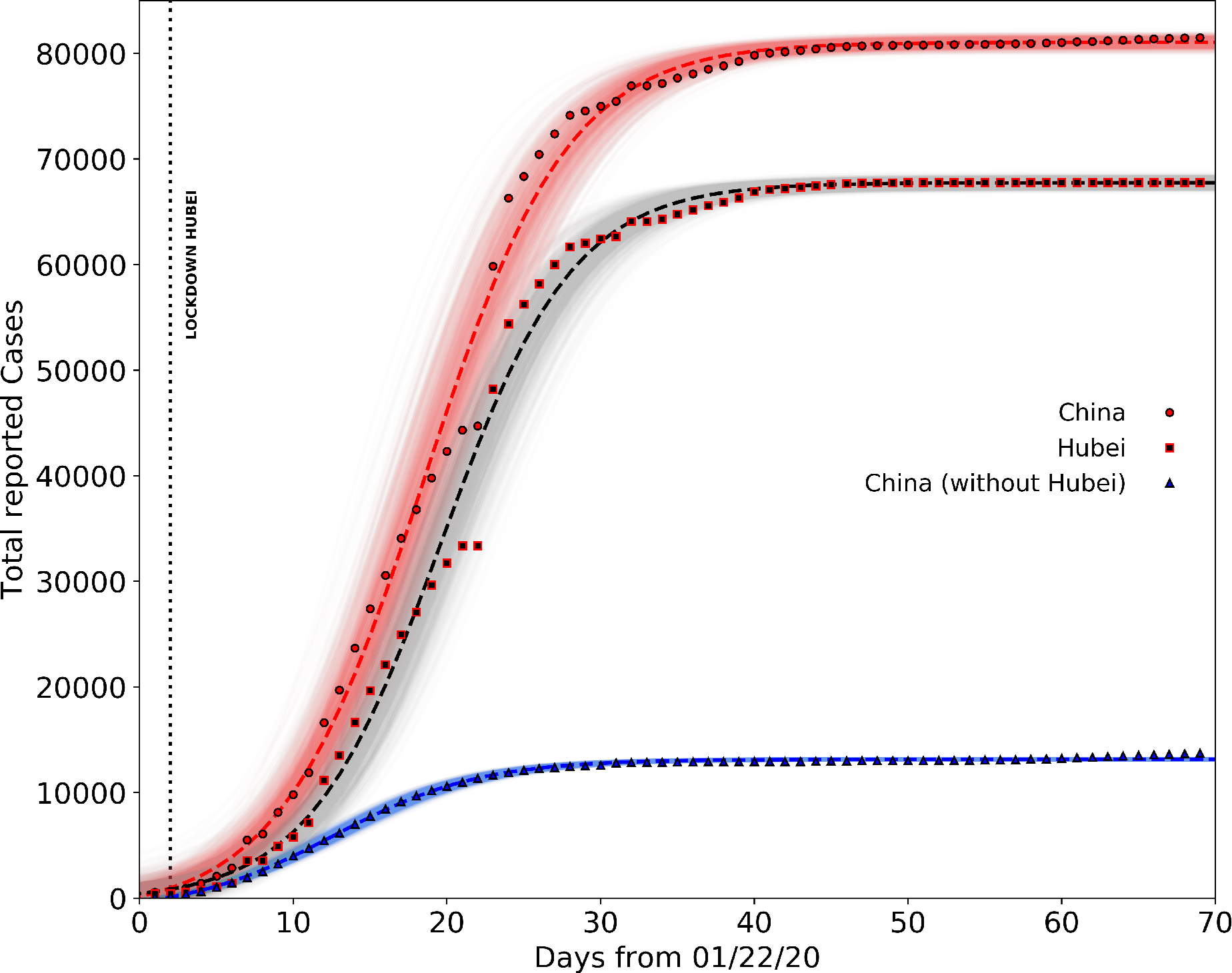
Total Covid-19 cases reported in China from January, 22nd to February, 25th according to Johns Hopkins University repository. Circles, squares and triangles represent total Covid-19 cases registered in China, region of Hubei and China without Hubei, respectively while red, black and blue dashed lines are the associated logistic fits. Shaded areas represent the family of curves obtainable by making the fit parameters vary within their confidence intervals. Fit parameters: Logistic(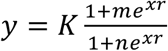;*China*:*K* = (811± 3) · 10^2^,*m*= −0.9 ±0.6,*n*=53 ±9,*r*=0.214± 0.008); *Hubei*:*K* = 678 ± 3,*m*= −0.4±1.2,*n*= 100 ± 20,*r*=0.232 ±0.01); *China without Hubei*:*K* =(131.6 ±0.3) · 10^3^,*m*= −1.4 ±0.11,*n*= 14±1.4,*r*=0.208 ±0.006).

To describe the evolution of the epidemic in Italy, as we explained, we decided to use the cumulative daily number of deceases to fit the logistic function, simply because is a much more reliable quantity when compared to the total reported cases. Figure 3a-b shows the fits of the deceased data reported in Italy (green), Lombardia (blue) and Italy without Lombardia (red), until March 30^th^, with a logistic function and its derivative. From the derivative of the logistic function, we observe that the model gives a rather good fit to the data until March 26^th^, whereas, after such a date, the number of deceases remains high, so perturbing the data around the peak, which does not decrease as the curve would previously predict. However, if we separate the data of Lombardia from the rest of Italy we see that the result is much more regular (red line). The curves, logistic and its derivative, are in this case both well constrained, and the peak value seems to have occurred on March 27^th^. The anomaly of the data of Lombardia is even more evident by considering the data of Emilia Romagna Region, which is the second more affected region according to the official reports. Figure 4a-b shows in red the logistic fit (and its derivative) to the cumulative (and new) number of deaceases in Emila Romagna. Also in this case, it is evident the data are well fitted and the curves well constrained. The peak value occurs around March 24^th^, coherently with this region being among the first ones where the infection began. Besides Emilia-Romagna, which represents a good example of a region in which the epidemic is already ‘mature’ (in the sense that the peak of deceases has already been overcome), it is interesting to see what happens in one of the Southern Italy regions, for example Calabria, where the infection presumibly started later. This data is also shown in Figure 4 for comparison, in blue. It is clear that, since the epidemic here has not reached yet the peak value, the curves are poorly constrained and the future evolution is still very uncertain. The difference between Emilia Romagna and Calabria curves illustrates well the different behaviour of the epidemic in the different Regions, and mainly between the Northern and the Southern ones.

**Figure 3.**
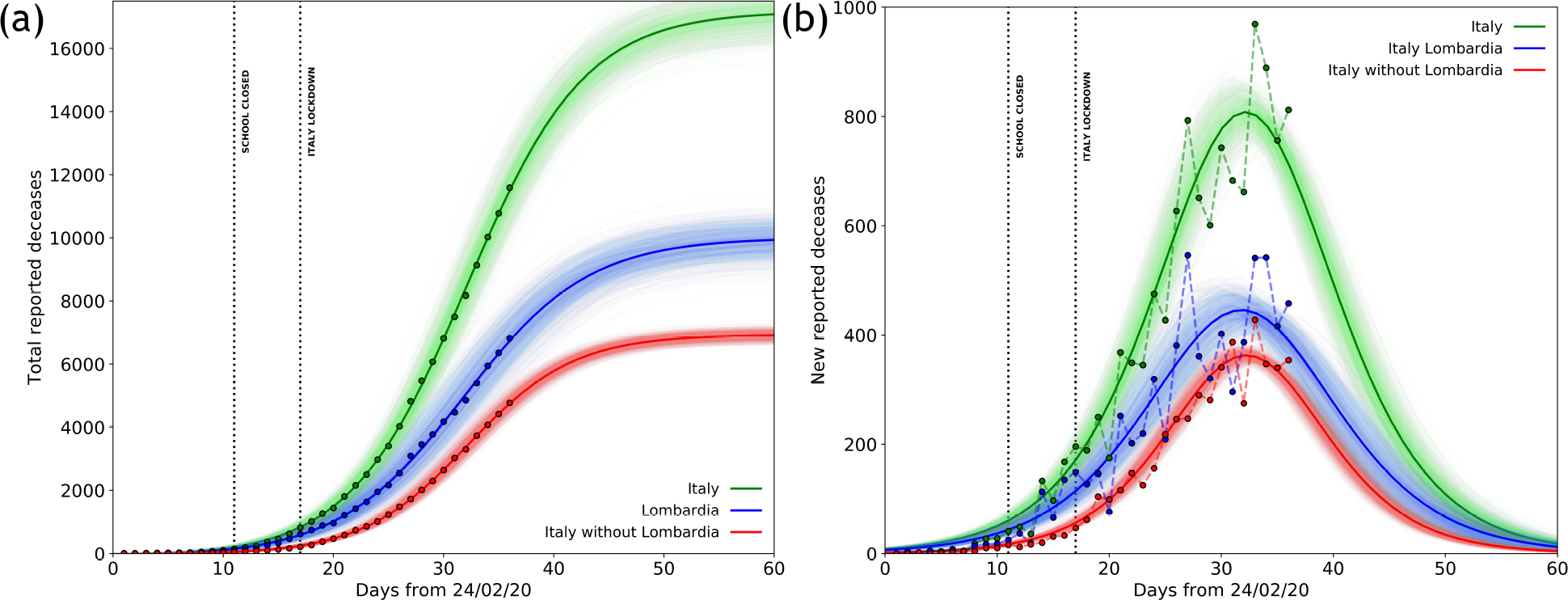
Deceases reported by Italian Civil Protection. a) Total deceases reported in Italy (green dots), Lombardia (blue dots) and Italy without Lombardia (red dots). from February, 24^th^ to March, 30^th^ and the corresponding logistic fit in solid lines. Dotted vertical lines mark the dates of Italian school lockdown and Italy total lockdown. b) Same as in a) using the new reported deceases and fitting the derivate of the logistic curve. Shaded areas represent the family of curves obtainable by making the fit parameters vary within their confidence intervals. Fit parameters: Logistic(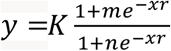;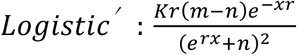;*Italy*:*K* = (178 ±4) · 10^2^,*m*= −3.1 ±0.5,*n* = 380 ±30,*r*=0.183 ±0.004); *Lombardia*:*K* = 100 ±4 · 10^2^,*m*= −2.8 ±0.5,*n* = 270 ±30,*r*=0.176 ±0.006);*ItalywithoutLombardia*:*K* = (74± 2) · 10^2^,*m*= −4.0 ±0.8,*n*= 740± 60,*r*=0.201 ±0.004).

**Figure 4.**
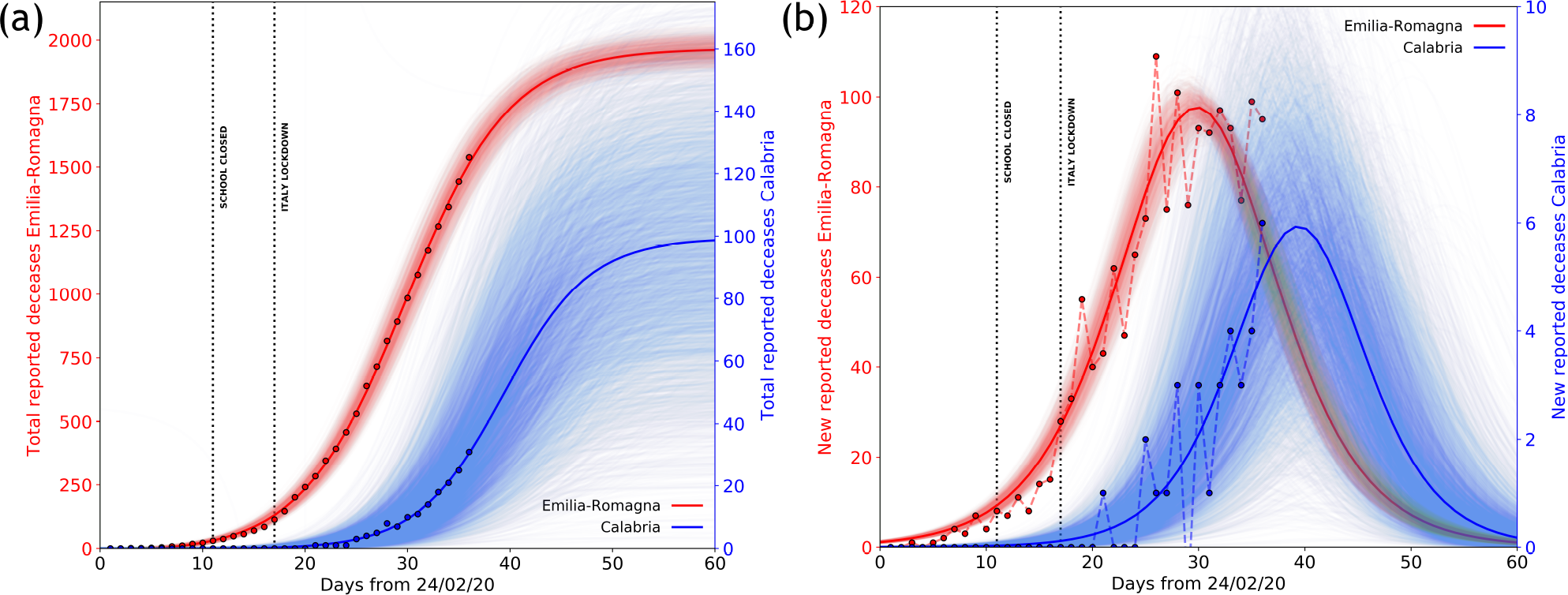
Deceases reported by Italian Civil Protection for Emilia Romagna and Calabria regions. a) Total, cumulative deceases reported in Emilia Romagna (red dots) and in Calabria (blue dots) from February, 24^th^ to March, 30^th^ according to Italian Civil Protection and the corresponding logistic fit obtained from the data. Dotted vertical lines mark the dates of Italian school lockdown and Italy total lockdown. b) Same as in a) using the new reported deceases and fitting the derivate of the logistic curve. Shaded areas represent the family of curves obtainable by making the fit parameters vary within their confidence intervals. Fit parameters: Logistic (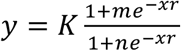;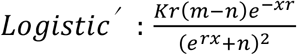;;*Emilia* −*Romagna*:*K* =1970 ±40,*m*= −3.0 ±0.6,*n*= 360 ±30,*r*=0.197 ±0.004); *Calabria*:*K* = 100 ± 20,*m*= −30 ±13,*n*=(12 ± 3) · 10^3^,*r*=0.208 ±0.006).

To estimate now the cumulative number of infections, we decided to use several tentative values of IFR: the one computed from the University of Oxford, IFR=0.2%; the one computed from the mortality on the Diamond Princess, IFR=1.3%; and one much larger, in case the IFR for Italy would be much larger than in other countries, IFR=5%. This high percetange can take into account the hospital saturation issues that could have locally amplified the IFR. Such a large range of IFRs for conversion is useful to check the final amount of total infected but also to determine the portion of unreported cases in respect to the official numbers.

In order to estimate the total number of infected cases, we take the less-noisy data from Italy without Lombardia but we normalize the number of deceases to be equal to the total Italian deceases (including the Lombardia ones) at the last date (March 30^th^). This procedure is used in order to remove some data scattering introduced by the Lombardia reported number of deceases without, on the other hand, underestimating the total number of infected. In Figure 5, we report the fit of the estimated amount of total cases in whole Italy with the procedure that we just described. The different logistic curves (blue, red and green), present in the figure, are the estimated number of contagious people for the different IFR values of 0.2%, 1.3% and 5%, respectively. From these values we also subtract the number of cases reported by Italian Civil Protection to estimate the number of unreported contagious cases (light blue, pink, light green); the parameters obtained from the best fits are reported in Table 2. It is evident that the number of reported cases is only a minor fraction of the estimated number of contagious people. In particular, for an IFR of 0.2%, 1.3% and 5% we estimate in the whole Italy a total number of cases at March 30^th^ (35 days from the first declared red zone) of about 8 milions, 1.2 milions and 300000, depending on the 0.2, 1.3 and 5 % IFR value, respectively. With a total number of reported cases of about 100000, Italy might be strongly underestimating the total number of infected people (including asymptomatic and pauci- symptomatic persons) of 98.75%, 91.6% or 66.6% depending on the IFR. From the time dependence of the logistic function, we observe that the inflections of the respective curves are now exceeded by the data points, whatever the conversion factor IFR is used. This probably means that Italy as a whole have already overcome the maximum number of new daily infections. In particular, the peak value of the infections should have been reached around March 11^th^, one week after the closing of schools and just after the lockdown of Italy. Moreover, our results predict that the point of 95% of the maximum value of the best fitting logistic curves has been already reached on the last week of March, and that, within the first week of April, the real number of contagious people will be already well within the saturation, so that it practically will not increase anymore.

**Table 2.** Logistic fit 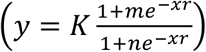 parameters for the estimated (total and undetected) Covid-19 cases in Italy based on three different IFR hypotheses: 0.2% (blue and light blue dots), 1.3% (red and pink dots) and 5% (green and light green dots) (see Fig. 5).

| **Total estimated cases (IFR 0.2 %)** |  |  |  |
| --- | --- | --- | --- |
| *K: $(81 \pm 1.6) \cdot 10^5$* | *m: $-3.7 \pm 0.7$* | *q: $860 \pm 60$* | *r: $0.211 \pm 0.003$* |
| **Total undetected cases (IFR 0.2 %)** |  |  |  |
| $K: (80 \pm 1.6) \cdot 10^5$ | $m: -3.7 \pm 0.7$ | $q: 860 \pm 60$ | $r: 0.211 \pm 0.003$ |
| **Total estimated cases (IFR 1.3 %)** |  |  |  |
| $K: (137 \pm 3) \cdot 10^4$ | $m: -3.1 \pm 0.5$ | $n: 380 \pm 30$ | $r: 0.184 \pm 0.004$ |
| **Total undetected cases (IFR 1.3 %)** |  |  |  |
| $K: (132 \pm 3) \cdot 10^4$ | $m: -3.1 \pm 0.5$ | $n: 380 \pm 30$ | $r: 0.183 \pm 0.004$ |
| **Total estimated cases (IFR 5 %)** |  |  |  |
| $K: (357 \pm 9) \cdot 10^3$ | $m: -3.1 \pm 0.5$ | $n: 380 \pm 30$ | $r: 0.183 \pm 0.004$ |
| **Total undetected cases (IFR 5 %)** |  |  |  |
| $K: (302 \pm 6) \cdot 10^3$ | $m: -3.0 \pm 0.5$ | $n: 370 \pm 30$ | $r: 0.188 \pm 0.004$ |

**Figure 5.**
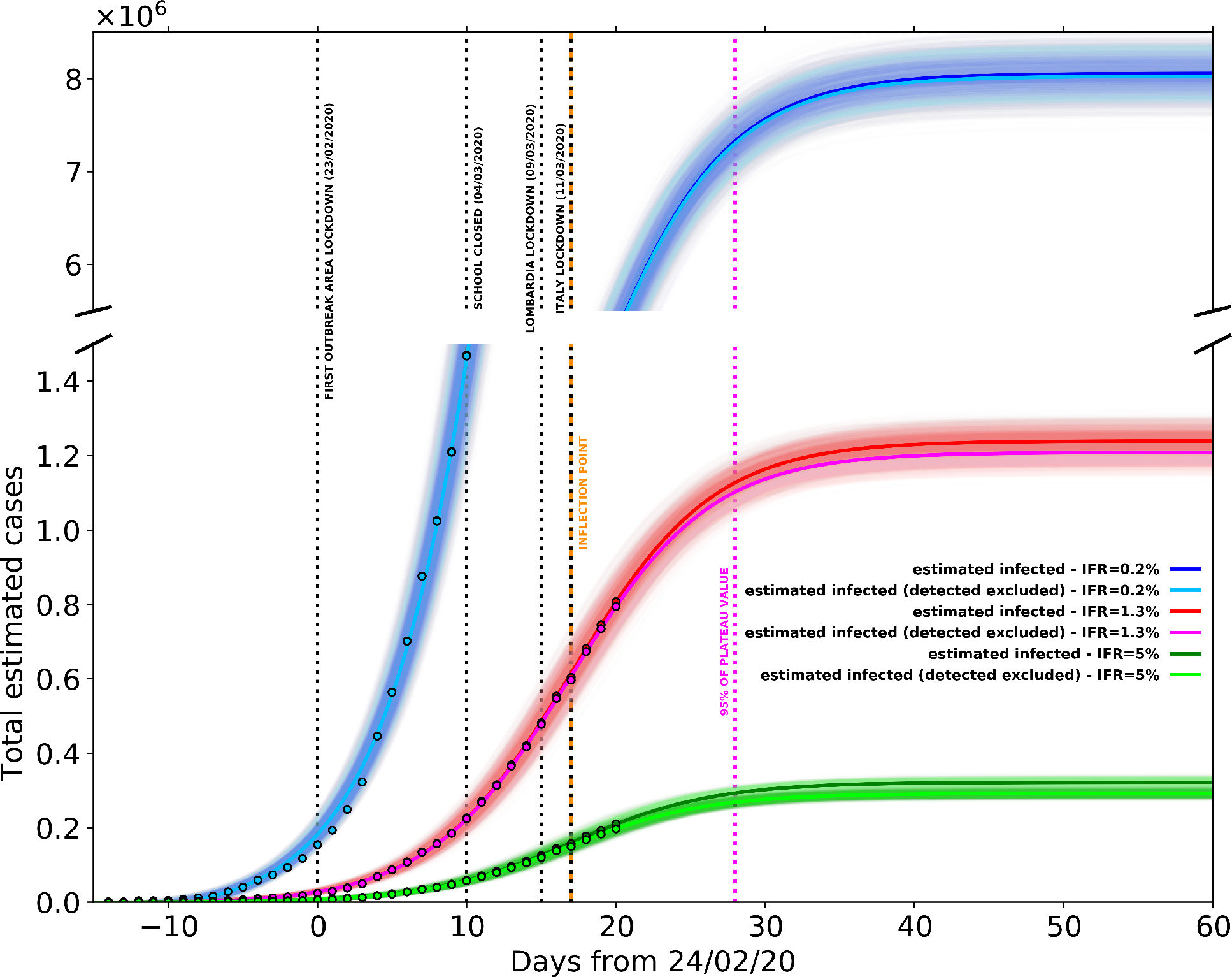
Estimated (total and undetected) Covid-19 cases in Italy based on three different IFR hypotheses: 0.2% (blue and light blue dots), 1.3% (red and pink dots) and 5% (green and light green dots). Blue and sky blue solid lines represent logistic fits of total and undetected estimated cases with IFR=0.2%, respectively. Red and pink solid lines represent logistic fits of total and undetected estimated cases with IFR=1.3%, respectively. Green and light green solid lines represent logistic fits of total and undetected estimated cases with IFR=5%, respectively. Black dotted vertical lines mark the dates of Codogno area lockdown, Italian schools’ lockdown, Lombardia lockdown and Italy lockdown. Dark orange dashed vertical line marks the inflection points of the three curves representing the total infected estimates; magenta dotted vertical line marks the 95% of the plateau of the three curves. Shaded areas represent the family of curves obtainable by making the fit parameters vary within their confidence intervals. Best fitting parameters are listed in Table 2.

It is possible that after the epidemic spread in Italy will be over, growth functions different from the logistic like Gompertz (1832), Janoschek (1957) or Richards (1959) sigmoids could be more suited to describe the epidemic spread because they allow their derivative to be non-symmetric and therefore can result in a more generalized integral function. A non symmetric integral curve, with a slower derivative in the descending part after the peak, could result for instance by a differential shift of epidemic curves for different Regions (and particularly for the Southern ones), due to the delay of the infection starting with respect to Lombardia, the first nucleus of infection spreading. When compared to the logistic function, a typical variation of these more generalized growth functions is the shift of the inflection point a few days later and the increase of the saturation value. Because here we are highlighting an extreme underestimation of the reported cases in Italy already with a simple logistic function, our main result would be even more inflated by the use of other growth functions. Nevertheless, despite this inherent limitation of the logistic function, as of March, 30^th^, data which overcome the inflexion point are reasonably well constrained, also if they do not cover the higher part of the curve. This, together with the test on the infection curve of China already shown in Figure 2, demonstrates that the logistic curve provides a robust approximation of the epidemic, at least during its spread.

## Discussion

The Covid-19 epidemic in Italy shows very abnormal features, in terms of severity of disease and, particularly, mortality. The CFR (defined as the ratio between the number of deceased and the number of recorded infected) reaches here very high values, disproportioned with respect to any other country: about 11%, rising to about 16% in the Lombardia Region, which alone represents half of the total number of infected people in Italy. Estimating a reliable value of the IFR (the ratio between the number of deceases and the total number of infected people) during the epidemic, for a virus hard to diagnose and requiring complex laboratory procedures, is very difficult. In this case, we considered two approaches: the first one, used by University of Oxford (Oke and Henegan, 2020) is based on the minimum reliable CFR values. The other one benefits of the possibility to study a very peculiar laboratory-like case study, namely the case of Diamond Princess, the cruise ship remaining at anchor in Japan (in the port of Yokohama) in which all the passengers and crew (3771 people) were tested for Covid-19. University of Oxford assumed a 50% of non recorded infected; in this way, they computed a value of IFR=0.2%. In the alternative approach, we considered that in the Diamond Princess cruise ship 712 infected were detected, and 9 of them died. So, the mostly unbiased value of IFR=CFR=1.3% can be computed. We have then considered these values as the limiting values defining the range of IFR. If we assume that the infection number underestimation is the only reason for the large overestimation of the mortality rate, in order to determine the real number of infected cases able to correct the apparent mortality to the true value, we should multiply the ‘official’ number of recorded infections by the ratio between the CFR=11.3% and the IFR (=0.2% or 1.3%). In this way, we obtain a real number of infected people in Italy (today, March 30^th^) ranging between about 885000 and 5.7 million people infected.

Even if this number appears very high (mainly with respect to the total number of reported cases of less than 90.000 in China, where however, using the same assumption, this amount should be multiplied by 4), it is not unreasonable for several reasons. Firstly, besides the high number of tests made in Italy (about 500000), this is still only a small fraction of the total population (60 million people), and does not take into account the number of asymptomatic and pauci-symptomatic cases. This is confirmed by observing that CFR in Italy is very similar to percent of death detected in hospital case series (Chen at al., 2020). The large number of potential daily infections in Italy during mobility for work or study can be also independently estimated by the statistics of the use of public transportation (the most likely to cause infection due to the high number of people assembled in small space). It results that 30 million people moves in Italy each day for work or study, with peak values in Lombardia (ISTAT report, 2017): about 56% of these people use public transportation; two thirds of the total travel for more than 15 minutes. So, considering only such a number of assembled people, and neglecting the likely high incidence of student assemblage in schools, of people in work places, and the more or less occasional frequentations of restaurants, clubs, pubs, sport matches, theatres, supermarkets etc., we get a minimum estimate of about 11 million people staying in close contact each day. A final indication that such number of effective infected cases is not unrealistic comes from the exponential fit of the first days of infection (Fig. 1); by extrapolating that exponential curve till March 30^th^ it would predict about 1 million infected cases. Obviously, such an hypothesis would assume that the exponential increase lasted till March 30^th^ (or however till few days before) and that the real exponential curve was missed because the limited number of tests progressively sampled a smaller and smaller percentage of the true daily number. From any point of view, in conclusion, a real number of 0.9-5.7 ×10^6^ infected people today does not appear unrealistic.

Other reasons could however equally affect the very high mortality observed (as CFR): discarding the existence in Italy of a virus strain significantly different and more aggressive (hypothesis actually impossible to verify because there is not yet the Italian genome available), a possible contribution could come from the relative high air pollution (from particulate matter PM10, PM2.5 and Ozone) of the zone around the Po Valley, where more than 50% of counted infections are clustered. Among the other hypothesized causes, the high average age (however lower than Japan, which showed a much lower CFR) is not likely to play a fundamental role; nor it is the number of smoking people (lower than the EU average). The observed high antibiotic-resistance, the highest one in Europe, has been also indicated as a possible contributing factor. However, although such problem causes in Italy about one third (10000 cases; the double of France, and fourfold of Germany) of the total deceases for antibiotic resistance in the whole EU (33000 cases), it does not appear to significantly affect the mortality (CFR) associated to seasonal flu (the most reliable comparison we can do, to infer the possible effect on Covid-19 mortality), which seems not significantly higher with respect to other European countries. We can compare, for instance, the number of deaths directly or indirectly associated to seasonal flu (the number of indirect deaths is much more significant in this case, because it is mostly linked to bacteria super-infections) in Italy and in Germany (where Covid-19 CFR is about 1%). The average yearly number of such deaths in Italy is around 8000 (ISS, 2020); in Germany, the deaths associated with the 2017-2018 flu, although very severe that year, totalled 25.000 (Koch Institute, 2019). Another issue that often comes out as a possible bias in the mortality estimates for Italy is the way of counting equally people dead ‘with’ or ‘by’ the infection. Actually, we do not think this question can seriously affect the data, provided only infected people dead for pulmonary collapse (or else for collapse of other organs following a serious pulmonary disease) are counted, as likely. On the contrary, an interesting issue, given the high selectivity of the disease towards old and/or multi-pathology people, will be to understand, at the end of the year, the total deaths caused, cumulatively, by all the seasonal diseases (flu, infections from antibiotic-resistant bacteria, walking pneumonia, Covid-19).

A significant contribution to the increase of the “local” IFR could, on the contrary, have come from the saturation of the public Hospitals, and in particular from the limited number (as compared for instance to Germany) of ICU. Italy, before the Covid-19 epidemic, had in fact 8.4 ICU per 10^5^ citizens (5090 ICU total), whereas Germany had 34 of them (28000 ICU total). Also, some wrong actions taken by the public Hospitals in managing the first days of infection (testified by many media and by a dramatic percentage of infected medical staff), could have significantly enhanced the infection. As of March 30^th^, 2020, 51 doctors died and 6414 people from the medical staff have been infected (ISS daily Info, March 30^th^, 2020). Moreover, the anomalous behaviour (with a persisting high number of deceases) in Lombardia, whereas the trend of deceases for the whole Italy excluding Lombardia is much more regular, also suggests some problems in that region, which has the most critical numbers and where the infection started. Such problems would likely involve insufficience of the sanitary system, stressed above the critical point, by the extreme numbers of people hospitalized (11883) or in ICUs (1324).

In this paper, we also use an indirect procedure, based on the analysis of the cumulative number of deceases which is the only reliable datum, in order to forecast the short-term evolution of the epidemic in Italy. As we have discussed, the number of recorded infections is not statistically reliable for such analyses, since inhomogeneous in time and in space (among different Regions). Considering correctly the average time shift of the death with respect to the infection (16 days used here) we show that decease data, converted to infection data using a large range of possible IFR, are well fitted by logistic functions, all of them indicating the time of 95% of total infection (i.e. the starting of the flat part of the function, which represents the end of infection), is expected around the last days of March. The point of inflection of the logistic best fitting functions, which corresponds to the peak of the infections in Italy, occurred around March 11^th^ given the interval of confidence over the delay time from sympthomps to death (Jung et al 2020). Looking at Figure 5, this date occurs just after the lockdown of the whole Italy and it demonstrates the high mitigating impact of the lockdowns; however, since it also occurs only from 4 to 10 days after the closing of the schools, it probably indicates that this measure has also been effective in containing the infection. Indeed, the high effectiveness of the closure of schools for containing epidemic spread has been specifically assessed by a number of epidemiologic studies (i.e. Adda, 2016).

Our results also put in evidence the shift of the infections curves between the Northern regions where the infection started (i.e. Emilia Romagna) and the Southern Italy regions (i.e. Calabria) where the peak is not reached yet, and therefore the fitting curves are still unconstrained and the future trends very uncertain. These results, based on the trend of the number of casualties, further confirm that the infection data we record today are only dependent on the variable and inhomogeneous sampling and have nothing to do with the true statistical evolution of the epidemic. Looking at these very inhomogeneous data, it is very clear that, since several days, the increase or decrease of new daily infections just depend on the number of tests. This is an obvious consequence of the fact, demonstrated here, that the true number of infections is much larger than the small sample tested: so, more tests you do, more cases you record. The information that the epidemic in the whole Italy is reaching the saturation (and hopefully end, as forecasted, within the first half of April), although not apparent from the tested number of new infections, should be seriously considered in order to decide how to tune and eventually release the next measures for the containment of the infection.

## Conclusions

We analysed the Covid-19 epidemic in Italy, which showed to be the largest and most lethal in the world. We discussed the causes of such anomalous behaviour of the disease in Italy, where mortality appears much higher than in other countries. Among the various hypotheses made till now to explain such an anomalous behaviour, it appears that neither the old age of population, nor the observed antibiotic-resistance of population (by far the highest one in Europe), nor even the smoking level, should have significant effects on the observed mortality. The most reasonable effect would involve a strong underestimation of the extension of the infection (implying a factor 8.7 to 56.5 more cases than the tested ones). A possible, further contribution could be given by the very high fine powder (and Ozone) pollution (one of the highest ones in Europe), which on one hand could facilitate the virus transmission, on the other hand could make more vulnerable and stressed the lung, causing heavier damages upon impact with virus. Another factor emerging from our analyses (as well as from media) that likely amplified mortality, could have been the scarse preparedness and possible initial faults of the sanitary system, mainly in Lombardia where the epidemic first blew up in few days.

The problem of underestimation of the number of infections, coupled with a non homogeneous sampling and testing of the positive cases, makes these data unreliable to use for statistics aimed to forecast the epidemic evolution. We then use here the cumulative daily number of deceases, corrected for an appropriate IFR, to simulate the evolution of the epidemic by a logistic function. When corrected for IFR and time shift between infection and death, these data can be well fitted by a logistic function, and show that actually the peak of infection has been overcome more than two weeks ago, and the saturation of the curve (end of epidemic) is expected within the first week of April or few days later. For the Southern Italy regions, the saturation of the curve will be most- likely delayed of a few days. The information here obtained about the possible evolution of the epidemic and its likely end, which is not at all evident from the very inhomogeneous sampling of infections actually performed, should be seriously considered in order to decide what measures to undertake or relax in the next future. Moreover, considering the likely effectiveness of the school’s closure, which is probably the minimally invasive measure in a social and economic sense, it should be considered as a primary measure when the lockdown will end. The study of this unprecedented medical catastrophe will hopefully give robust indications to avoid, in the future, the same errors and understatements. It will also help to seriously consider the need to improve health system both in hospital and primary health care settings, whose capability should be considered invaluable, also using a purely economic criterion.

## Data Availability

All the data are taken by public datasets, referenced in the paper. They are free to be used for anyone.

